# Associations of Healthy Lifestyle and Socioeconomic Status with Cognitive Function in U.S. Older Adults

**DOI:** 10.1101/2023.01.24.23284961

**Authors:** Xin Wang, Kelly M. Bakulski, Henry L. Paulson, Roger L. Albin, Sung Kyun Park

**Affiliations:** Department of Epidemiology, School of Public Health, University of Michigan, Ann Arbor, Michigan, USA, 48109; Michigan Alzheimer’s Disease Center, University of Michigan, Ann Arbor, Michigan, USA, 48109; Department of Neurology, University of Michigan, Ann Arbor, Michigan, USA, 48109; Neurology Service & GRECC, VAAAHS, Ann Arbor, Michigan, USA, 48109; Department of Environmental Health Sciences, School of Public Health, University of Michigan, Ann Arbor, Michigan, USA, 48109

**Author notes:** Correspondence to Xin Wang, Department of Epidemiology, University of Michigan, 6630 SPH I, 1415 Washington Heights, Ann Arbor, Michigan, USA, 48109-2029.

**Keywords:** socioeconomic status, healthy lifestyle, cognitive function, older adults

## Abstract

We investigated the complex relations of socioeconomic status (SES) and healthy lifestyles with cognitive functions among older adults in 1,313 participants, aged 60 years and older, from the National Health and Nutrition Examination Survey 2011-2014. Cognitive function was measured using an average of the standardized z-scores of the Consortium to Establish a Registry for Alzheimer’s Disease Word Learning and delayed recall tests, the Animal Fluency Test, and the Digit Symbol Substitution Test. Latent class analysis of family income, education, occupation, health insurance, and food security was used to define composite SES (low, medium, high). A healthy lifestyle score was calculated based on smoking, alcohol consumption, physical activity, and the Healthy-Eating-Index-2015. In survey-weighted multivariable linear regressions, participants with 3 or 4 healthy behaviors had 0.07 (95% CI: 0.005, 0.14) standard deviation higher composite cognitive z-score, relative to those with one or no healthy behavior. Participants with high SES had 0.37 (95% CI: 0.29, 0.46) standard deviation higher composite cognitive z-score than those with low SES. No statistically significant interaction was observed between healthy lifestyle score and SES. Our findings suggested that higher healthy lifestyle scores and higher SES were associated with better cognitive function among older adults in the United States.

## 1. Introduction

Dementia currently affects approximately 50 million people worldwide, and this number is expected to rise to 152 million by 2050.^1^ Maintaining cognitive function is crucial for promoting the health of the aging population. Individuals with low socioeconomic status (SES) are more likely to experience impaired cognitive function.^2–6^ SES has also been associated with functional and structural neural differences in wide range of cortical areas.^7^ Despite increasing life expectancy, there are widening SES inequalities in health.^8^ At this time, treatments for dementia are inadequate.^9^ Identifying susceptible populations and modifiable risk factors for cognitive decline are priorities for public health intervention.^10^ Comprehensive strategies are warranted to identify vulnerable groups who may benefit most from preventive interventions to reduce cognitive declines and socioeconomic disparities.

Most emphasis is now directed towards individual-level interventions on potentially modifiable lifestyle factors.^11^ Diets rich in fruit and vegetables, abstinence from smoking, and regular exercise are associated with better cognitive performance and lower risk of dementia.^12–14^ Despite extensive investigations, clear understanding of the roles of healthy lifestyles and SES in cognitive health is still lacking.^15^ Few attempts have been made to investigate associations between combined lifestyle factors and cognitive function. For studies focusing on SES, most previous studies used single socioeconomic variables (e.g., education, occupation, income) to represent SES.^16,17^ SES is a complex, multifactorial construct, and composite SES measurements reflecting multiple socioeconomic factors are needed to characterize.^18,19^ Furthermore, evidence suggests that healthy lifestyles could play a role in health outcomes patterned by SES, particularly cardiometabolic disease and mortality;^15^ yet, it is unknown whether healthy lifestyles could alleviate socioeconomic inequalities in cognitive health.

We used data from the National Health and Nutrition Examination Survey (NHANES) to examine the complex relationships of SES and lifestyle factors with cognitive function in older adults. We defined individual-level SES with a composite of education, occupation, poverty-to-income ratio, health insurance, and food security. A composite healthy lifestyle score was constructed based on health behaviors including smoking status, alcohol consumption, physical activity, and diet. We hypothesized that higher composite SES and healthy lifestyle scores were associated with better cognitive function in a cross-sectional sample of U.S. older adults.

## 2. Results

### 2.1. Univariate analyses: participant characteristics and cognitive function by SES and healthy lifestyle score

The analytic sample had a mean age of 69.3 years. LCA was used to classify SES based on multiple factors (**Table S2**). The high SES class was characterized by income-to-poverty ratio ≥4, white collar occupations, college education or higher, health insurance, and full food security. The medium SES class was characterized by income-to-poverty ratio >1 and <4, blue collar occupation, and food security. The low SES class was characterized by income-to-poverty ratio <4, blue collar occupations, less than high school education, government insurance or uninsured, and food insecurity. Within the sample, 581 were of high SES, 351 were of medium SES, and 381 were of low SES. Low SES participants tended to be women, non-White people, not married, born outside of the United States, have less alcohol consumption, and less healthy diets (**Table 1**). For healthy lifestyles, 341 study participants had 0 or 1 healthy behavior, 512 had 2 healthy behaviors, and 460 had 3 or 4 healthy behaviors (**Table 2**). Participants with less healthy behaviors were more likely to be non-White people, born in the United States, and have blue collar occupations.

**Table 1.**
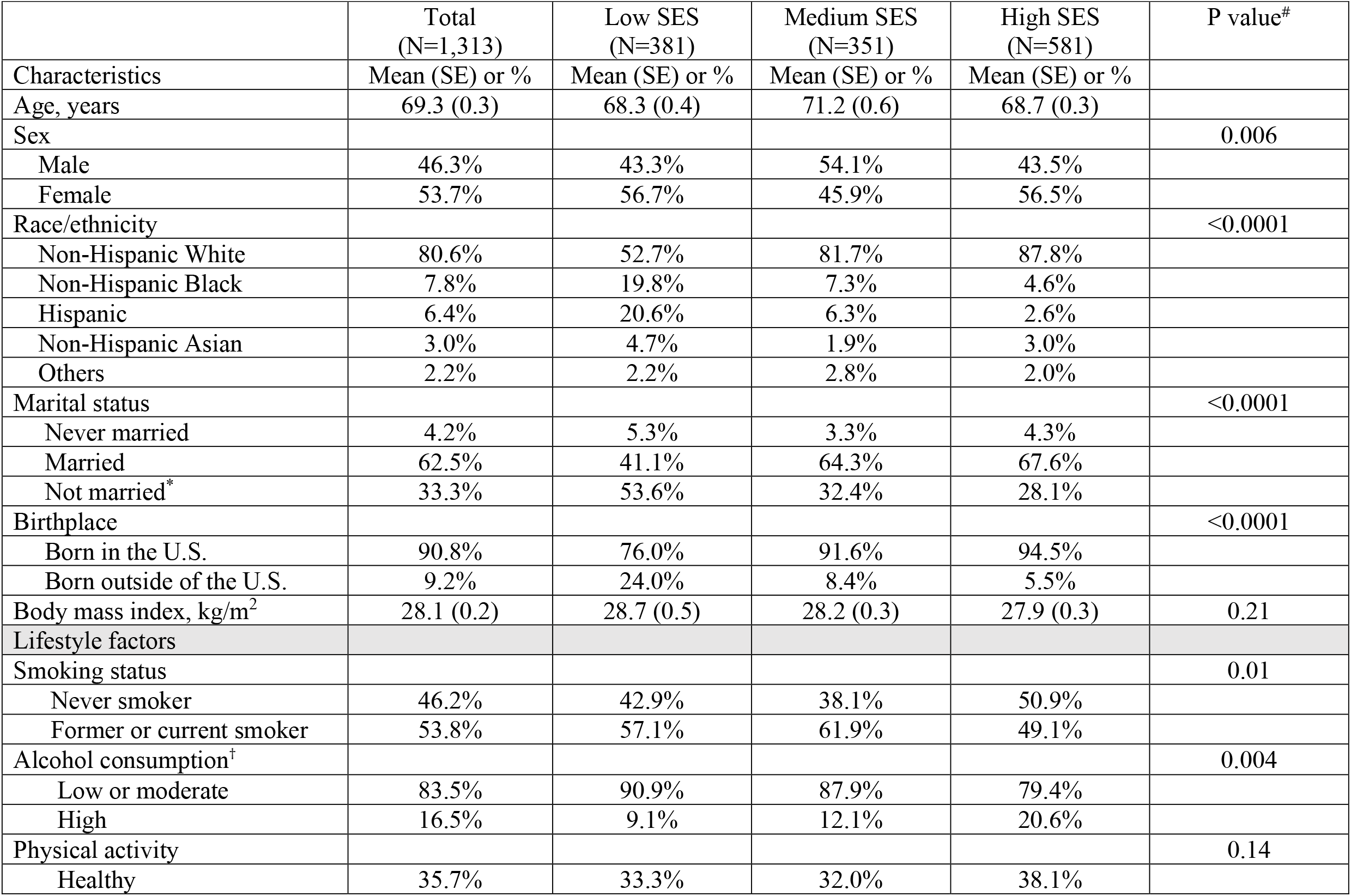
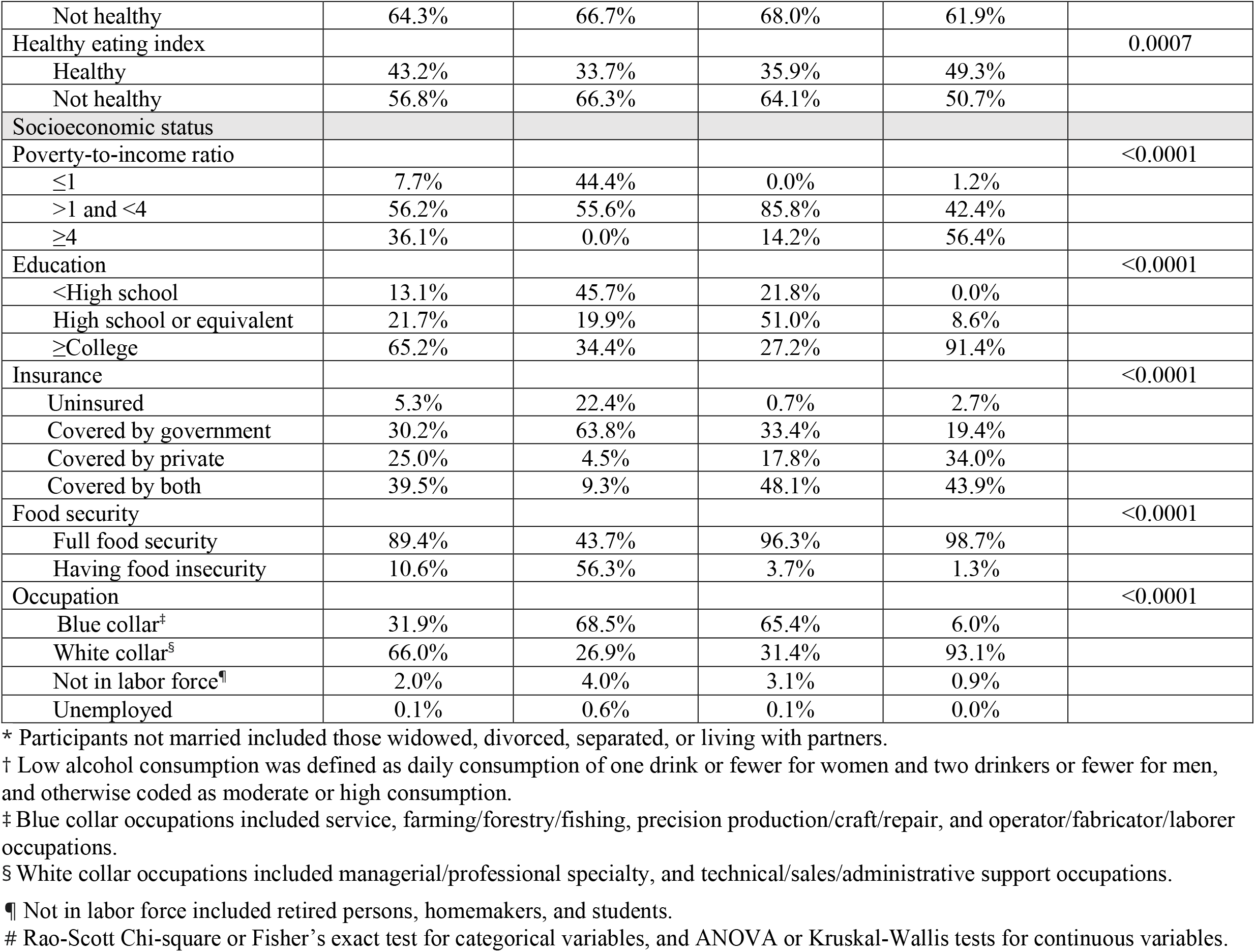
Survey-weighted participant characteristics by socioeconomic status (SES: low, medium, and high) from the U.S. National Health and Nutrition Examination Survey 2011-2014.

|  | Total<br>(N=1,313) | Low SES<br>(N=381) | Medium SES<br>(N=351) | High SES<br>(N=581) | P value<sup>#</sup> |
| --- | --- | --- | --- | --- | --- |
| Characteristics | Mean (SE) or % | Mean (SE) or % | Mean (SE) or % | Mean (SE) or % |  |
| Age, years | 69.3 (0.3) | 68.3 (0.4) | 71.2 (0.6) | 68.7 (0.3) |  |
| Sex |  |  |  |  | 0.006 |
| Male | 46.3% | 43.3% | 54.1% | 43.5% |  |
| Female | 53.7% | 56.7% | 45.9% | 56.5% |  |
| Race/ethnicity |  |  |  |  | <0.0001 |
| Non-Hispanic White | 80.6% | 52.7% | 81.7% | 87.8% |  |
| Non-Hispanic Black | 7.8% | 19.8% | 7.3% | 4.6% |  |
| Hispanic | 6.4% | 20.6% | 6.3% | 2.6% |  |
| Non-Hispanic Asian | 3.0% | 4.7% | 1.9% | 3.0% |  |
| Others | 2.2% | 2.2% | 2.8% | 2.0% |  |
| Marital status |  |  |  |  | <0.0001 |
| Never married | 4.2% | 5.3% | 3.3% | 4.3% |  |
| Married | 62.5% | 41.1% | 64.3% | 67.6% |  |
| Not married* | 33.3% | 53.6% | 32.4% | 28.1% |  |
| Birthplace |  |  |  |  | <0.0001 |
| Born in the U.S. | 90.8% | 76.0% | 91.6% | 94.5% |  |
| Born outside of the U.S. | 9.2% | 24.0% | 8.4% | 5.5% |  |
| Body mass index, kg/m<sup>2</sup> | 28.1 (0.2) | 28.7 (0.5) | 28.2 (0.3) | 27.9 (0.3) | 0.21 |
| Lifestyle factors |  |  |  |  |  |
| Smoking status |  |  |  |  | 0.01 |
| Never smoker | 46.2% | 42.9% | 38.1% | 50.9% |  |
| Former or current smoker | 53.8% | 57.1% | 61.9% | 49.1% |  |
| Alcohol consumption<sup>†</sup> |  |  |  |  | 0.004 |
| Low or moderate | 83.5% | 90.9% | 87.9% | 79.4% |  |
| High | 16.5% | 9.1% | 12.1% | 20.6% |  |
| Physical activity |  |  |  |  | 0.14 |
| Healthy | 35.7% | 33.3% | 32.0% | 38.1% |  |
| Not healthy | 64.3% | 66.7% | 68.0% | 61.9% |  |
| Healthy eating index |  |  |  |  | 0.0007 |
| Healthy | 43.2% | 33.7% | 35.9% | 49.3% |  |
| Not healthy | 56.8% | 66.3% | 64.1% | 50.7% |  |
| Socioeconomic status |  |  |  |  |  |
| Poverty-to-income ratio |  |  |  |  | <0.0001 |
| ≤1 | 7.7% | 44.4% | 0.0% | 1.2% |  |
| >1 and <4 | 56.2% | 55.6% | 85.8% | 42.4% |  |
| ≥4 | 36.1% | 0.0% | 14.2% | 56.4% |  |
| Education |  |  |  |  | <0.0001 |
| <High school | 13.1% | 45.7% | 21.8% | 0.0% |  |
| High school or equivalent | 21.7% | 19.9% | 51.0% | 8.6% |  |
| ≥College | 65.2% | 34.4% | 27.2% | 91.4% |  |
| Insurance |  |  |  |  | <0.0001 |
| Uninsured | 5.3% | 22.4% | 0.7% | 2.7% |  |
| Covered by government | 30.2% | 63.8% | 33.4% | 19.4% |  |
| Covered by private | 25.0% | 4.5% | 17.8% | 34.0% |  |
| Covered by both | 39.5% | 9.3% | 48.1% | 43.9% |  |
| Food security |  |  |  |  | <0.0001 |
| Full food security | 89.4% | 43.7% | 96.3% | 98.7% |  |
| Having food insecurity | 10.6% | 56.3% | 3.7% | 1.3% |  |
| Occupation |  |  |  |  | <0.0001 |
| Blue collar<sup>‡</sup> | 31.9% | 68.5% | 65.4% | 6.0% |  |
| White collar<sup>§</sup> | 66.0% | 26.9% | 31.4% | 93.1% |  |
| Not in labor force<sup>¶</sup> | 2.0% | 4.0% | 3.1% | 0.9% |  |
| Unemployed | 0.1% | 0.6% | 0.1% | 0.0% |  |

\* Participants not married included those widowed, divorced, separated, or living with partners.
† Low alcohol consumption was defined as daily consumption of one drink or fewer for women and two drinkers or fewer for men, and otherwise coded as moderate or high consumption.
‡ Blue collar occupations included service, farming/forestry/fishing, precision production/craft/repair, and operator/fabricator/laborer occupations.
§ White collar occupations included managerial/professional specialty, and technical/sales/administrative support occupations.
¶ Not in labor force included retired persons, homemakers, and students.

**Table 2.**
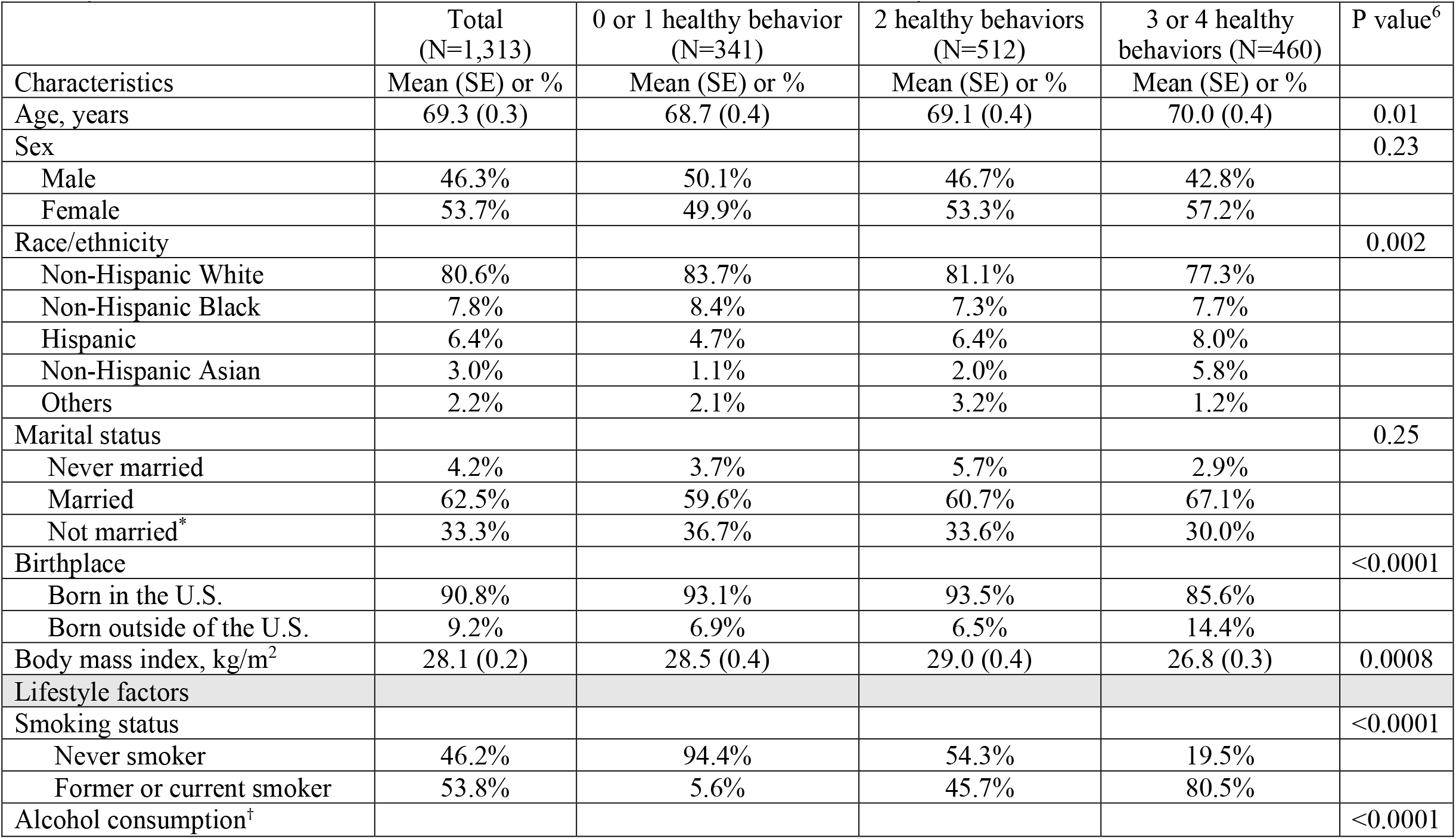
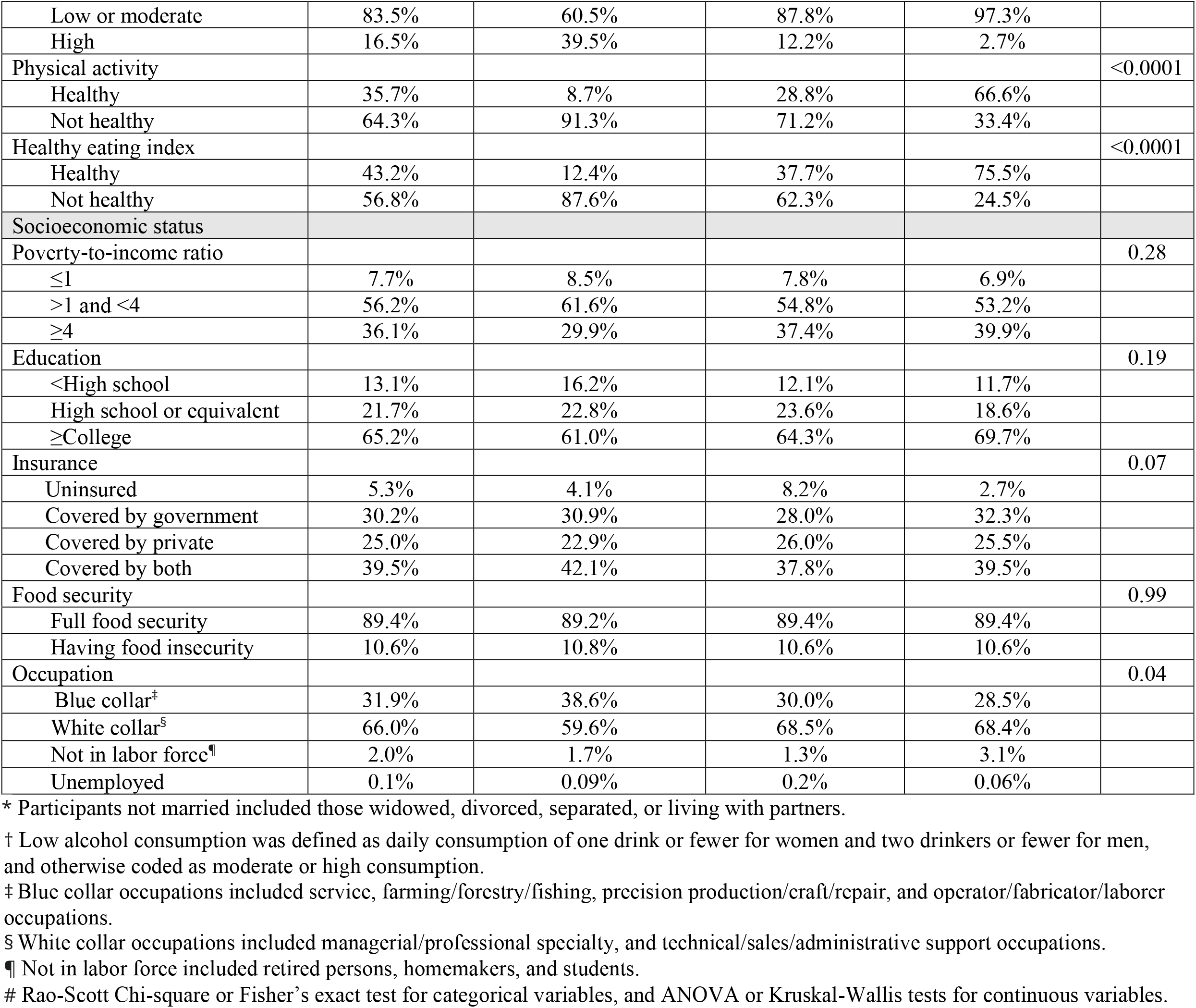
Survey-weighted participant characteristics by healthy lifestyle score (HLS: 0 or 1 healthy behavior; 2 healthy behaviors; 3 or 4 healthy behaviors) from the U.S. National Health and Nutrition Examination Survey 2011-2014.

|  | Total<br>(N=1,313) | 0 or 1 healthy behavior<br>(N=341) | 2 healthy behaviors<br>(N=512) | 3 or 4 healthy<br>behaviors (N=460) | P value<sup>6</sup> |
| --- | --- | --- | --- | --- | --- |
| Characteristics | Mean (SE) or % | Mean (SE) or % | Mean (SE) or % | Mean (SE) or % |  |
| Age, years | 69.3 (0.3) | 68.7 (0.4) | 69.1 (0.4) | 70.0 (0.4) | 0.01 |
| Sex |  |  |  |  | 0.23 |
| Male | 46.3% | 50.1% | 46.7% | 42.8% |  |
| Female | 53.7% | 49.9% | 53.3% | 57.2% |  |
| Race/ethnicity |  |  |  |  | 0.002 |
| Non-Hispanic White | 80.6% | 83.7% | 81.1% | 77.3% |  |
| Non-Hispanic Black | 7.8% | 8.4% | 7.3% | 7.7% |  |
| Hispanic | 6.4% | 4.7% | 6.4% | 8.0% |  |
| Non-Hispanic Asian | 3.0% | 1.1% | 2.0% | 5.8% |  |
| Others | 2.2% | 2.1% | 3.2% | 1.2% |  |
| Marital status |  |  |  |  | 0.25 |
| Never married | 4.2% | 3.7% | 5.7% | 2.9% |  |
| Married | 62.5% | 59.6% | 60.7% | 67.1% |  |
| Not married* | 33.3% | 36.7% | 33.6% | 30.0% |  |
| Birthplace |  |  |  |  | <0.0001 |
| Born in the U.S. | 90.8% | 93.1% | 93.5% | 85.6% |  |
| Born outside of the U.S. | 9.2% | 6.9% | 6.5% | 14.4% |  |
| Body mass index, kg/m<sup>2</sup> | 28.1 (0.2) | 28.5 (0.4) | 29.0 (0.4) | 26.8 (0.3) | 0.0008 |
| Lifestyle factors |  |  |  |  |  |
| Smoking status |  |  |  |  | <0.0001 |
| Never smoker | 46.2% | 94.4% | 54.3% | 19.5% |  |
| Former or current smoker | 53.8% | 5.6% | 45.7% | 80.5% |  |
| Alcohol consumption<sup>†</sup> |  |  |  |  | <0.0001 |
| Low or moderate | 83.5% | 60.5% | 87.8% | 97.3% |  |
| High | 16.5% | 39.5% | 12.2% | 2.7% |  |
| Physical activity |  |  |  |  | <0.0001 |
| Healthy | 35.7% | 8.7% | 28.8% | 66.6% |  |
| Not healthy | 64.3% | 91.3% | 71.2% | 33.4% |  |
| Healthy eating index |  |  |  |  | <0.0001 |
| Healthy | 43.2% | 12.4% | 37.7% | 75.5% |  |
| Not healthy | 56.8% | 87.6% | 62.3% | 24.5% |  |
| Socioeconomic status |  |  |  |  |  |
| Poverty-to-income ratio |  |  |  |  | 0.28 |
| ≤1 | 7.7% | 8.5% | 7.8% | 6.9% |  |
| >1 and <4 | 56.2% | 61.6% | 54.8% | 53.2% |  |
| ≥4 | 36.1% | 29.9% | 37.4% | 39.9% |  |
| Education |  |  |  |  | 0.19 |
| <High school | 13.1% | 16.2% | 12.1% | 11.7% |  |
| High school or equivalent | 21.7% | 22.8% | 23.6% | 18.6% |  |
| ≥College | 65.2% | 61.0% | 64.3% | 69.7% |  |
| Insurance |  |  |  |  | 0.07 |
| Uninsured | 5.3% | 4.1% | 8.2% | 2.7% |  |
| Covered by government | 30.2% | 30.9% | 28.0% | 32.3% |  |
| Covered by private | 25.0% | 22.9% | 26.0% | 25.5% |  |
| Covered by both | 39.5% | 42.1% | 37.8% | 39.5% |  |
| Food security |  |  |  |  | 0.99 |
| Full food security | 89.4% | 89.2% | 89.4% | 89.4% |  |
| Having food insecurity | 10.6% | 10.8% | 10.6% | 10.6% |  |
| Occupation |  |  |  |  | 0.04 |
| Blue collar<sup>‡</sup> | 31.9% | 38.6% | 30.0% | 28.5% |  |
| White collar<sup>§</sup> | 66.0% | 59.6% | 68.5% | 68.4% |  |
| Not in labor force<sup>¶</sup> | 2.0% | 1.7% | 1.3% | 3.1% |  |
| Unemployed | 0.1% | 0.09% | 0.2% | 0.06% |  |

\* Participants not married included those widowed, divorced, separated, or living with partners.
† Low alcohol consumption was defined as daily consumption of one drink or fewer for women and two drinkers or fewer for men, and otherwise coded as moderate or high consumption.
‡ Blue collar occupations included service, farming/forestry/fishing, precision production/craft/repair, and operator/fabricator/laborer occupations.
§ White collar occupations included managerial/professional specialty, and technical/sales/administrative support occupations.
¶ Not in labor force included retired persons, homemakers, and students.

**Table 3** presents the distributions of the cognitive composite z-score and its components. Participants with higher SES, on average, had higher test scores of CERAD delayed recall, Animal Fluency, DSST, as well as the composite z-score, compared to participants with lower SES. The mean (SE) test scores were 6.7 (0.1) for CERAD delayed recall, 19.8 (0.3) for Animal Fluency, 58.8 (0.7) for DSST, and 0.41 (0.02) for the composite z-score among participants with high SES. In contrast, the mean (SE) test scores were 5.7 (0.2) for CERAD delayed recall, 16.2 (0.4) for Animal Fluency, 41.8 (1.5) for DSST, and -0.07 (0.04) for the composite z-score among participants with low SES.

**Table 3.**
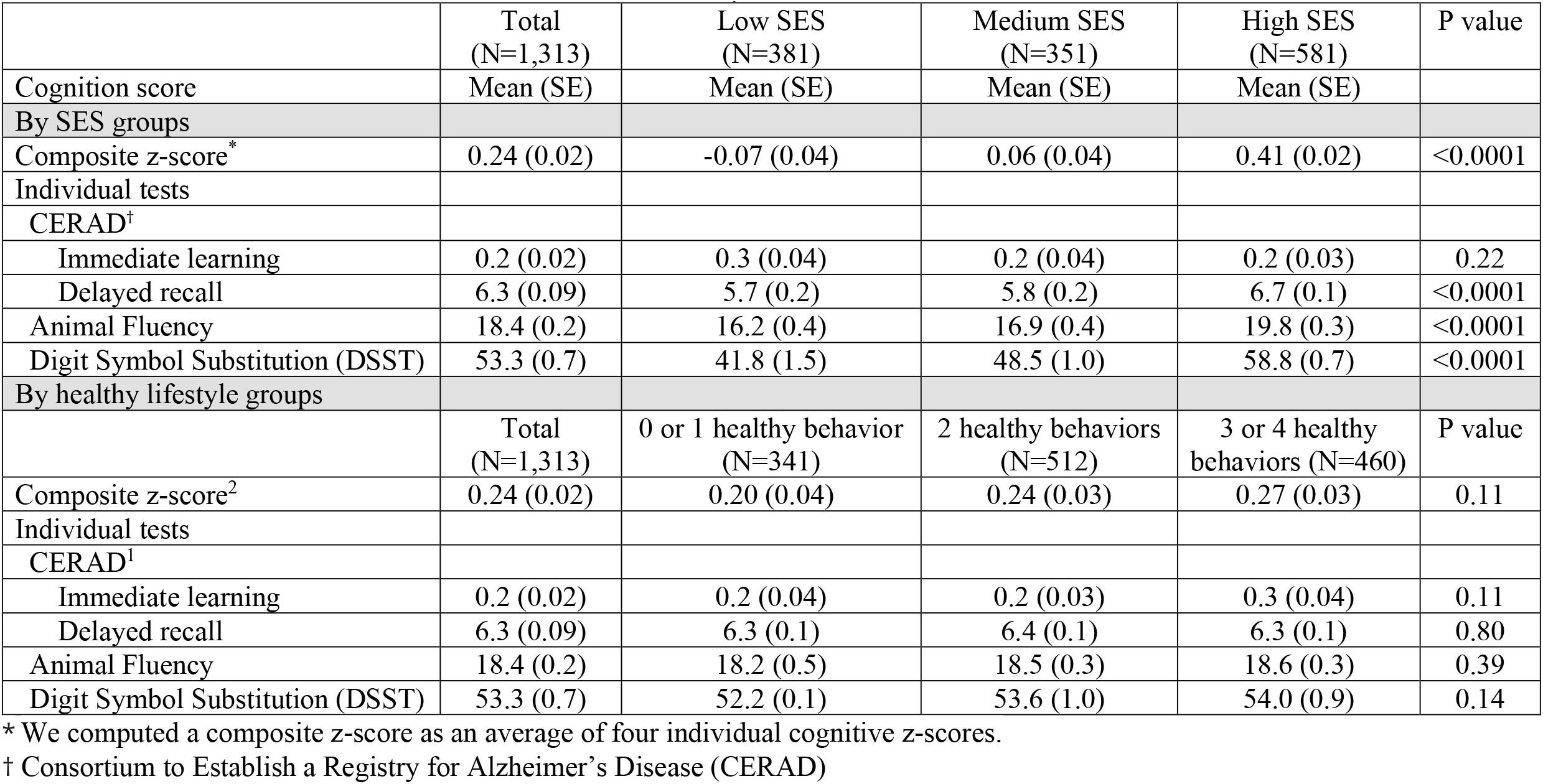
Survey-weighted cognitive composite score and its components by socioeconomic status (SES) and healthy lifestyle score from the U.S. National Health and Nutrition Examination Survey 2011-2014.

|  | Total<br>(N=1,313) | Low SES<br>(N=381) | Medium SES<br>(N=351) | High SES<br>(N=581) | P value |
| --- | --- | --- | --- | --- | --- |
| Cognition score | Mean (SE) | Mean (SE) | Mean (SE) | Mean (SE) |  |
| By SES groups |  |  |  |  |  |
| Composite z-score<sup>*</sup> | 0.24 (0.02) | -0.07 (0.04) | 0.06 (0.04) | 0.41 (0.02) | <0.0001 |
| Individual tests |  |  |  |  |  |
| CERAD<sup>†</sup> |  |  |  |  |  |
| Immediate learning | 0.2 (0.02) | 0.3 (0.04) | 0.2 (0.04) | 0.2 (0.03) | 0.22 |
| Delayed recall | 6.3 (0.09) | 5.7 (0.2) | 5.8 (0.2) | 6.7 (0.1) | <0.0001 |
| Animal Fluency | 18.4 (0.2) | 16.2 (0.4) | 16.9 (0.4) | 19.8 (0.3) | <0.0001 |
| Digit Symbol Substitution (DSST) | 53.3 (0.7) | 41.8 (1.5) | 48.5 (1.0) | 58.8 (0.7) | <0.0001 |
| By healthy lifestyle groups |  |  |  |  |  |
|  | Total<br>(N=1,313) | 0 or 1 healthy behavior<br>(N=341) | 2 healthy behaviors<br>(N=512) | 3 or 4 healthy<br>behaviors (N=460) | P value |
| Composite z-score<sup>2</sup> | 0.24 (0.02) | 0.20 (0.04) | 0.24 (0.03) | 0.27 (0.03) | 0.11 |
| Individual tests |  |  |  |  |  |
| CERAD<sup>†</sup> |  |  |  |  |  |
| Immediate learning | 0.2 (0.02) | 0.2 (0.04) | 0.2 (0.03) | 0.3 (0.04) | 0.11 |
| Delayed recall | 6.3 (0.09) | 6.3 (0.1) | 6.4 (0.1) | 6.3 (0.1) | 0.80 |
| Animal Fluency | 18.4 (0.2) | 18.2 (0.5) | 18.5 (0.3) | 18.6 (0.3) | 0.39 |
| Digit Symbol Substitution (DSST) | 53.3 (0.7) | 52.2 (0.1) | 53.6 (1.0) | 54.0 (0.9) | 0.14 |

\* We computed a composite z-score as an average of four individual cognitive z-scores.
† Consortium to Establish a Registry for Alzheimer's Disease (CERAD)

### 2.2. Primary analysis: adjusted associations between composite exposure measures and cognitive score

To assess the associations between cumulative SES and lifestyle factors, we tested the adjusted relationships of healthy lifestyle score and composite SES with composite cognitive z-score (**Table 4**). Participants with 3 or 4 healthy behaviors, on average, had 0.07 (95% CI: 0.005, 0.14) standard deviation higher composite cognitive z-scores than those with one or less healthy behavior. Participants with high SES, on average, had 0.37 (95% CI: 0.29, 0.46) standard deviation higher composite cognitive z-scores than those with low SES.

**Table 4.**
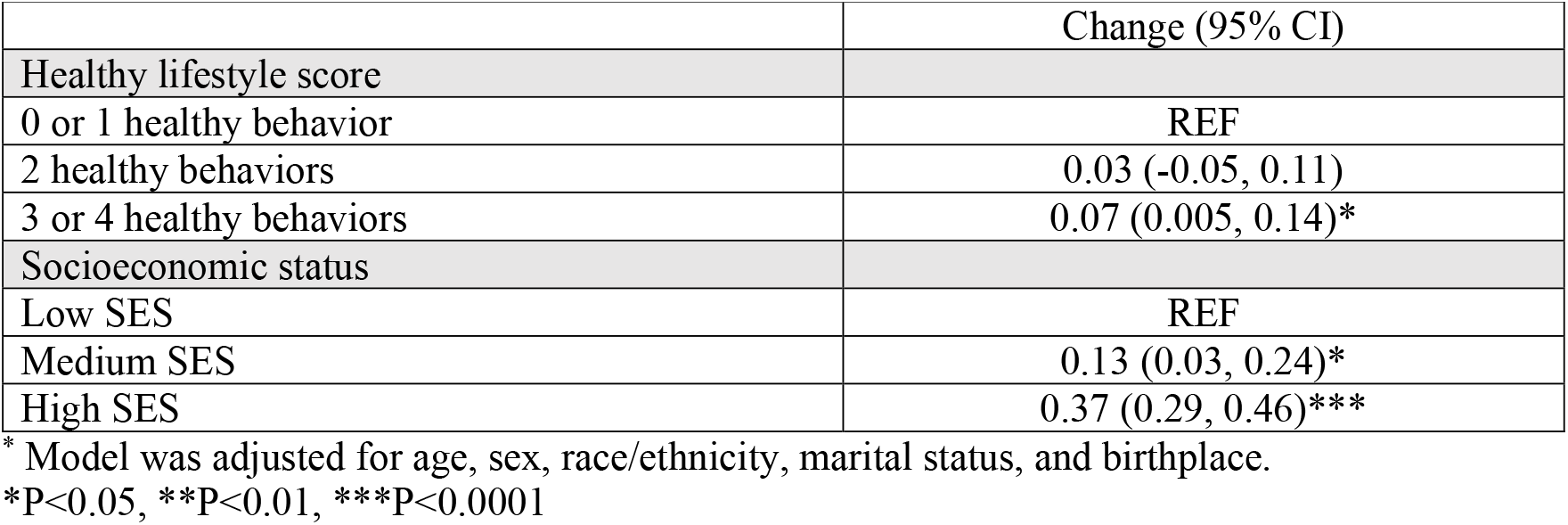
Survey-weighted differences and 95% confidence intervals (95% CIs)^*^ in cognitive composite z-score with socioeconomic status and healthy lifestyle from the U.S. National Health and Nutrition Examination Survey 2011-2014.

**Figure 1** presents the joint associations of lifestyle and SES with composite cognitive z-score, according to the combination of the nine categories of healthy lifestyle score and composite SES. The interaction between the healthy lifestyle score and composite SES was not statistically significant (P for interaction=0.06). Participants with high SES had better cognitive function compared to those with low SES, independent of the healthy lifestyle score.

**Figure 1.**
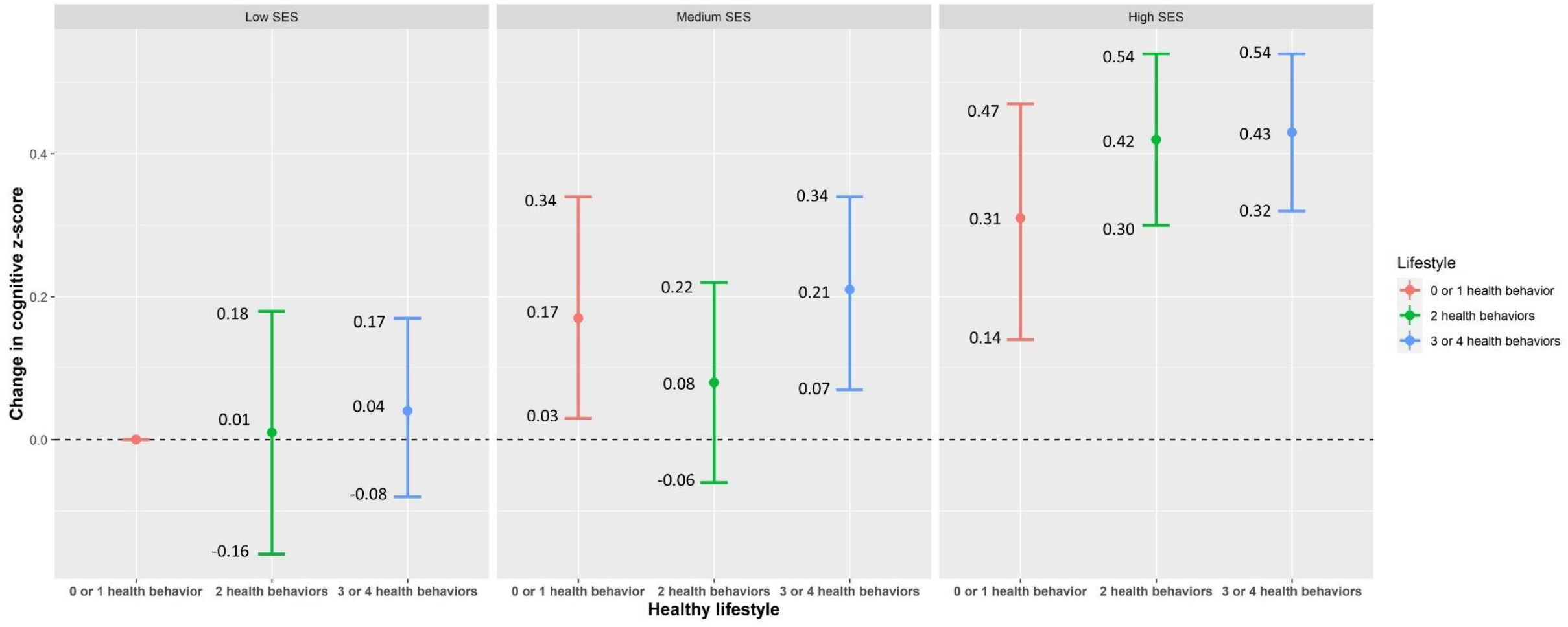
Survey-weighted changes in cognition z-score with an additional interaction term between socioeconomic status (SES) and healthy lifestyle from the U.S. National Health and Nutrition Examination Survey 2011-2014. Model was adjusted for age, sex, race/ethnicity, marital status, and birthplace. P for interaction between SES and healthy lifestyle was 0.06.

### 2.3. Secondary analysis: adjusted associations between individual exposure measures and cognitive score

We tested the associations between individual healthy lifestyle behaviors (mutually adjusted) and cognition score, adjusting for age, sex, race/ethnicity, marital status, and birthplace (Model 1, **Table 5**). On average, participants with low or moderate alcohol consumption had 0.11 (95% CI: -0.21, -0.02) standard deviation lower cognitive score compared to those with high alcohol consumption. Those with healthy physical activity levels had 0.08 (95% CI: 0.03, 0.14) standard deviation higher cognitive scores compared to those with unhealthy physical activity. Those with healthy eating had 0.08 (95% CI: 0.02, 0.15) standard deviation higher composite cognitive z-scores, compared to those with unhealthy eating habits.

**Table 5.**
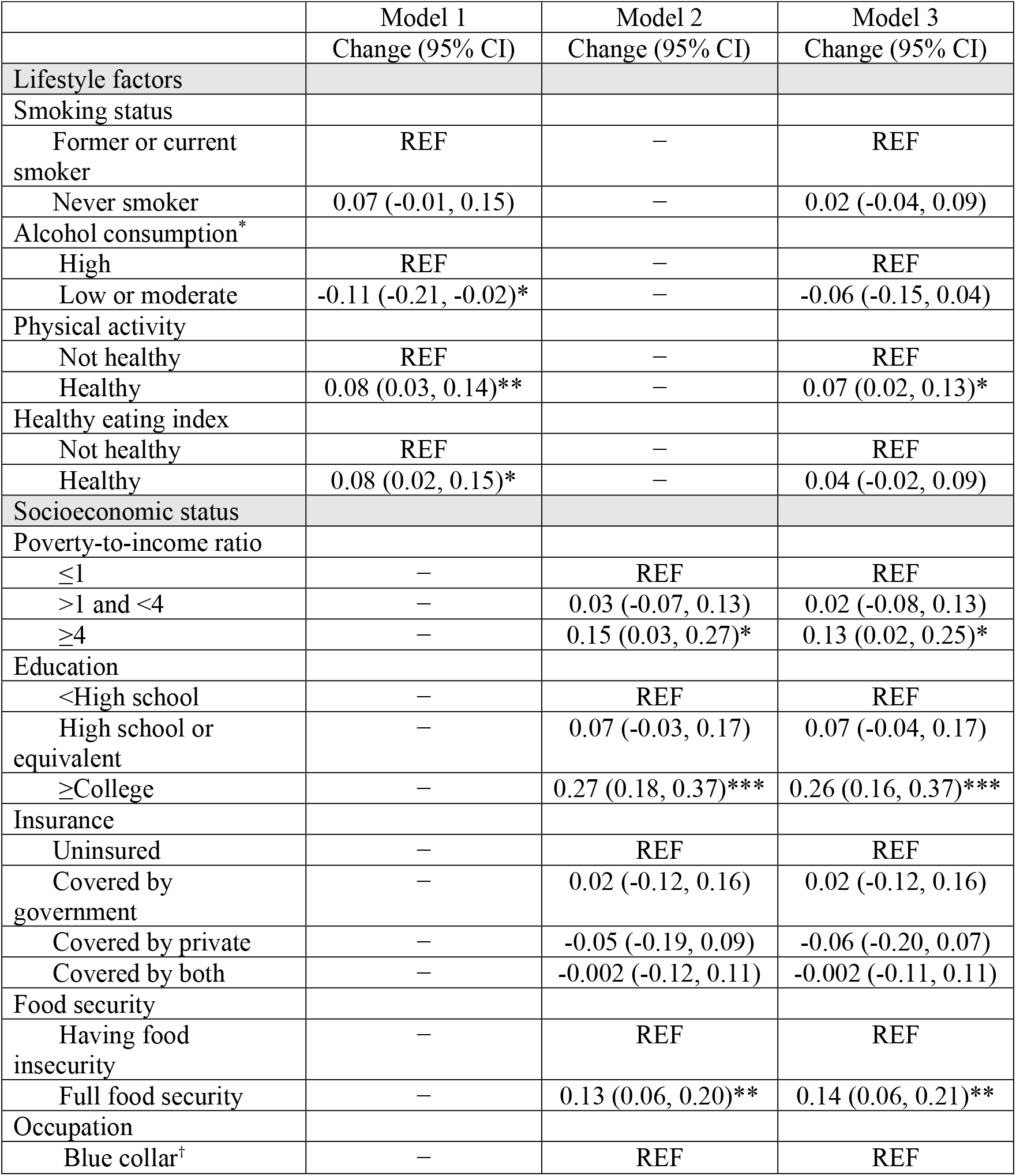
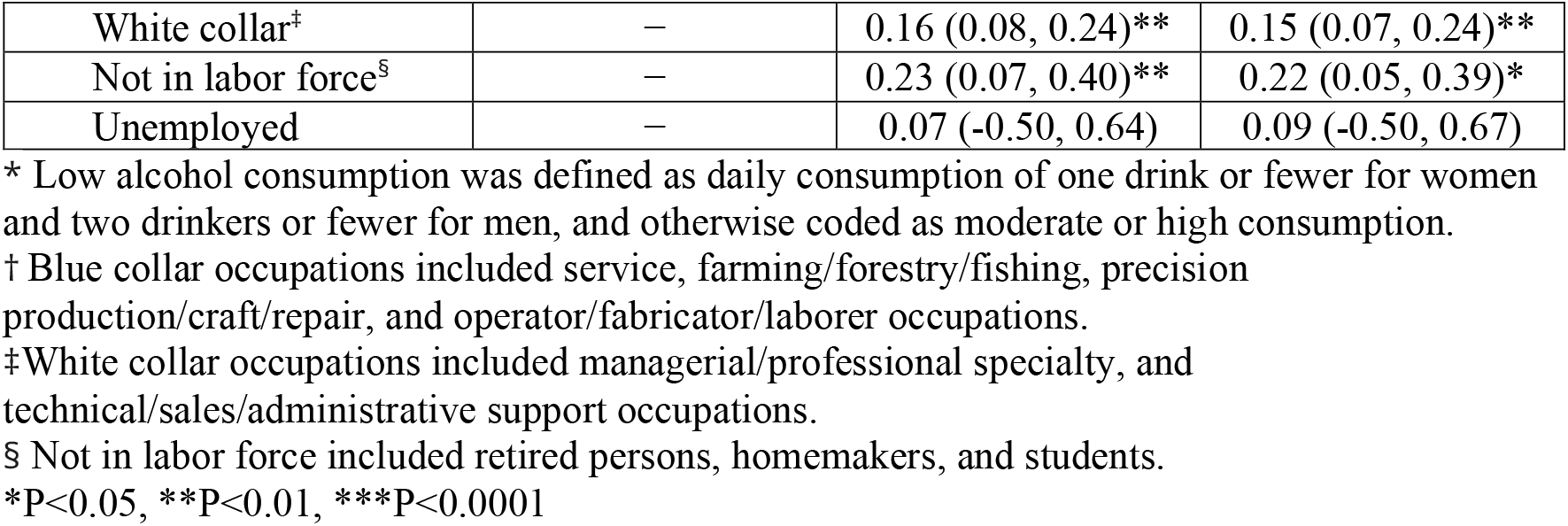
Survey-weighted differences and 95% confidence intervals (95% CIs) in cognitive composite z-score from linear regression models using data from the U.S. National Health and Nutrition Examination Survey 2011-2014.

|  | Model 1 | Model 2 | Model 3 |
| --- | --- | --- | --- |
|  | Change (95% CI) | Change (95% CI) | Change (95% CI) |
| **Lifestyle factors** |  |  |  |
| **Smoking status** |  |  |  |
| Former or current smoker | REF | — | REF |
| Never smoker | 0.07 (-0.01, 0.15) | — | 0.02 (-0.04, 0.09) |
| **Alcohol consumption\*** |  |  |  |
| High | REF | — | REF |
| Low or moderate | -0.11 (-0.21, -0.02)* | — | -0.06 (-0.15, 0.04) |
| **Physical activity** |  |  |  |
| Not healthy | REF | — | REF |
| Healthy | 0.08 (0.03, 0.14)** | — | 0.07 (0.02, 0.13)* |
| **Healthy eating index** |  |  |  |
| Not healthy | REF | — | REF |
| Healthy | 0.08 (0.02, 0.15)* | — | 0.04 (-0.02, 0.09) |
| **Socioeconomic status** |  |  |  |
| **Poverty-to-income ratio** |  |  |  |
| ≤1 | — | REF | REF |
| >1 and <4 | — | 0.03 (-0.07, 0.13) | 0.02 (-0.08, 0.13) |
| ≥4 | — | 0.15 (0.03, 0.27)* | 0.13 (0.02, 0.25)* |
| **Education** |  |  |  |
| <High school | — | REF | REF |
| High school or equivalent | — | 0.07 (-0.03, 0.17) | 0.07 (-0.04, 0.17) |
| ≥College | — | 0.27 (0.18, 0.37)*** | 0.26 (0.16, 0.37)*** |
| **Insurance** |  |  |  |
| Uninsured | — | REF | REF |
| Covered by government | — | 0.02 (-0.12, 0.16) | 0.02 (-0.12, 0.16) |
| Covered by private | — | -0.05 (-0.19, 0.09) | -0.06 (-0.20, 0.07) |
| Covered by both | — | -0.002 (-0.12, 0.11) | -0.002 (-0.11, 0.11) |
| **Food security** |  |  |  |
| Having food insecurity | — | REF | REF |
| Full food security | — | 0.13 (0.06, 0.20)** | 0.14 (0.06, 0.21)** |
| **Occupation** |  |  |  |
| Blue collar<sup>†</sup> | — | REF | REF |
| White collar<sup>‡</sup> | — | 0.16 (0.08, 0.24)** | 0.15 (0.07, 0.24)** |
| Not in labor force<sup>§</sup> | — | 0.23 (0.07, 0.40)** | 0.22 (0.05, 0.39)* |
| Unemployed | — | 0.07 (-0.50, 0.64) | 0.09 (-0.50, 0.67) |

\* Low alcohol consumption was defined as daily consumption of one drink or fewer for women and two drinkers or fewer for men, and otherwise coded as moderate or high consumption.
† Blue collar occupations included service, farming/forestry/fishing, precision production/craft/repair, and operator/fabricator/laborer occupations.
‡ White collar occupations included managerial/professional specialty, and technical/sales/administrative support occupations.
§ Not in labor force included retired persons, homemakers, and students.
\*P<0.05, \*\*P<0.01, \*\*\*P<0.0001

We tested the similarly adjusted associations between SES components and cognition score (Model 2, **Table 5**). We observed participants in the highest categories of family income (PIR ≥ 4) had 0.15 (95% CI: 0.03, 0.27) standard deviation higher composite cognitive scores, compared to those in the lowest categories (PIR ≤ 1 for family income). Participants in the highest category of education (college and above) had 0.26 (95% CI: 0.16, 0.37) standard deviation higher composite cognitive z-scores, compared to those in the less than high school education category. Participants with food security had 0.13 (95% CI: 0.06, 0.20) standard deviation higher composite cognitive z-scores than those with food insecurity. Participants who had white collar occupations had 0.16 (95% CI: 0.08, 0.24) standard deviation higher composite cognitive z-scores than those who had blue collar occupations.

We then included both lifestyle and SES components in similarly adjusted models of cognitive score (Model 3, **Table 5**). Similar associations were observed between all of SES components and cognitive z-score to the previous models without lifestyle component adjustment. However, after adjusting for SES components, physical activity was the only lifestyle factor still associated with cognitive z-score.

### 2.4. Sensitivity results

In sensitivity analyses of individual cognitive assessments, higher SES was associated with higher cognition measured with the CERAD immediate learning, Animal Fluency, and DSST (**Table S3**). When examining the associations using the weighted healthy lifestyle score, a similar positive association was observed (**Table S4**). Additional adjustment for BMI did not alter the associations (**Table S5**). In a subpopulation after excluding 283 participants with cardiovascular disease or stroke, similar associations of healthy lifestyle score and composite SES with composite cognitive z-score were observed (**Table S6** and **Table S7**). Similar associations were observed in the pooled analysis of 20 imputed datasets (**Table S8**).

## 3. Discussion

This study examined the associations of lifestyle and socioeconomic factors with cognitive function in a representative sample of adults aged 60 years and older in the United States. We summarized smoking status, alcohol consumption, physical activity, and healthy diet in a composite healthy lifestyle score and education, family income, occupation, health insurance, and food security into a composite SES. Both higher healthy lifestyle scores and higher SES scores were independently associated with better cognitive function. These associations persisted after controlling for age, sex, race, marital status, and birthplace. The magnitude of the SES effect was considerably greater than the magnitude of healthy lifestyle factors. In individual component analyses, participants with higher physical activity, higher education, higher family income, better health insurance, food security, and white-collar occupations had better cognitive function performance. This study is the first to investigate the complex relationships between SES, healthy lifestyles, and cognition.

Given population ageing and lack of therapies to halt the progression to dementia, identification of vulnerable populations and modifiable risk factors for age-related impaired cognitive function is of substantial public health importance. Lifestyle factors are potentially modifiable factors. The associations of various individual lifestyle factors such as healthy diet,^20^ physical activity,^21^ and smoking^22^ with cognitive function and dementia were described previously, which is consistent with our study results. Additionally, we evaluated the combinations of multiple lifestyle factors given that those factors are not independent of each other and the complex and multifactorial etiology of cognitive impairment and dementia. Three large trials examined the effects of multiple lifestyle interventions on cognitive function – the Finnish Geriatric Intervention Study to Prevent Cognitive Impairment and Disability (FINGER), the French Multidomain Alzheimer Preventive Trial (MAPT)^23^ and the Dutch Prevention of Dementia by Intensive Vascular Care (PreDIVA)^24^ trials – yielded inconsistent results. FINGER demonstrated some benefit, but MAPT and PreDIVA did not, though post-hoc analyses of participants at high dementia risk suggested benefits. Overall, our findings along with previous studies suggest that promoting a healthy lifestyle could be a feasible strategy that could have an impact on prevention for cognitive impairment and dementia. The associations between healthy lifestyles and cognitive function are biologically possible. Physical activity is associated with increased brain volume, elevated brain-derived neurotrophic factor levels, reduced psychological stress, reduced cardiometabolic risk factor levels, and enhanced amyloid beta clearance.^25^ Smoking exerts toxic effects through oxidative stress, neuroinflammation, and increase cardiometabolic risks.^25^ Healthy diets rich in nutrients and vitamins may ameliorate cognitive impairment through their effects on oxidative stress, inflammation, and cardiometabolic health.^25^

We found a larger effect of SES on cognitive function. Considerable evidence shows that SES components are associated with cognitive function. Higher education levels are consistently associated with higher cognitive function and lower risk of dementia.^5,6^ Lower income and sustained financial hardship were associated with lower cognitive function,^26,27^ and food insecurity adversely impacts the cognitive function.^28^ Studies examining the relationship between longest-held occupations and cognition found that blue collar occupations were associated with higher risks of cognitive impairment and Alzheimer’s disease.^29,30^ A large cohort study of U.S. found that low access to health care was associated with 25% higher odds of cognitive impairment.^31^ Our analysis observed similar associations between these single socioeconomic variables and cognitive function. More importantly, we constructed a composite SES variable given the nature of SES as a multidimensional construct comprising diverse socioeconomic factors, and that SES may affect health through multiple pathways.^32^ The association between composite SES and cognitive function suggests the socioeconomic inequalities in cognitive health in U.S. older adults, and thus, exploring strategies to reduce socioeconomic inequalities is needed.

The contribution of healthy lifestyles to the socioeconomic inequalities in health is widely discussed, particularly on cardiovascular outcomes and mortality. A recent systematic review estimated that lifestyles factors were responsible for approximately 20% of the socioeconomic inequalities in health, suggesting the promotion of healthy lifestyles since they may help alleviate socioeconomic inequalities in health.^15^ Contrary to these findings, we found that the healthy lifestyle score and SES were independently associated with cognitive function, though a borderline interaction between the healthy lifestyle score and SES was observed. Effect estimates of SES on cognitive function were also larger than those of the healthy lifestyle score. These findings may suggest that significant reductions in socioeconomic inequalities in health could not be achieved by only promoting healthy lifestyles, and additional modifiable factors such as environmental exposures, psychosocial factors, structural factors, and policies should be considered and evaluated. Some data suggests that SES inequality per se is a significant driver of health disparities. Lynch argues persuasively that more systemic interventions are required to address SES disparities in population health.^33^ Future research, especially prospective cohort studies or already performed lifestyle intervention trial datasets, are also needed to confirm our findings.

Strengths of the current study include examining the associations of healthy lifestyles and SES with cognitive function in a representative sample of U.S. older adults. In addition, we constructed a healthy lifestyle score and composite SES, acknowledging the complex and multifactorial impacts of lifestyle and socioeconomic factors on cognitive function. However, we acknowledge that the cross-sectional nature of NHANES data precludes the ability to assess longitudinal cognitive decline. We could not rule out the possibility of reverse causality that participants’ cognitive function could impact self-report on behavior factors such as diet. Additionally, both lifestyle and socioeconomic factors were measured in late life; thus, we are not able to investigate whether lifestyle and SES changes across the life course are associated with cognitive function at older ages. Therefore, it is critically important to examine the contribution of SES and lifestyle to cognition using the life course approach in the future. Finally, we were unable to completely eliminate residual confounding due to limited categorization of covariates, for example, lack of non-binary gender identity^34^ and *APOE* genotype^35^ due to the limitation in NHANES measures, despite we have controlled for many known confounders.

Our study provides evidence that higher SES, and healthy lifestyle are independently associated with a higher cognitive function performance in U.S. older adults. The magnitude of the SES association with cognitive function was considerably larger than the association between healthy lifestyle factors and cognitive performance. No statistically significant interaction between the healthy lifestyle and SES was observed, suggesting that reductions in socioeconomic inequalities in health cannot not be achieved by only promoting healthy lifestyles. Future studies with longitudinal design are needed to confirm our findings and explore more modifiable factors that help mitigate socioeconomic inequalities in health.

## 4. Methods

### 4.1. Study population

Data were from the NHANES, a large, nationally representative cross-sectional survey and physical examination conducted in 2-year cycles, assessing the health and nutritional status of the civilian, noninstitutionalized United States population. Details of the survey and laboratory procedures are published elsewhere.^36^ The comprehensive cognitive evaluation was available among 2,937 adults ≥ 60 years old from two continuous NHANES data releases: 1,364 from the 2011-2012 cycle and 1,573 from the 2013-2014 cycle. Our samples excluded 1,624 participants without information on lifestyle factors, SES factors, and covariates. Our final analytic sample included 1,313 participants from the NHANES 2011-2014, representative of 48,974,397 adults ≥ 60 years in the United States (**Figure S1)**.

### 4.2. Standard Protocol Approvals, Registrations, and Patient Consents

The NHANES protocol followed the ethical guidelines of the 1975 Declaration of Helsinki and was approved by the National Center for Health Statistics Research Ethics Review Board, and written informed consent was obtained from all participants or legally authorized representatives. The National Center for Health Statistics Research Ethnics Review Board approved all study protocols, and written informed consent was obtained from all participants. All methods were performed in accordance with relevant guidelines and regulations and followed the Strengthening the Reporting of Observational Studies for Epidemiology (STROBE) guidelines.^37^

### 4.3. Lifestyle factor assessment

Lifestyle factors consisted of multiple domains, including self-reported cigarette smoking, alcohol use, physical activity, and diet. Cigarette smoking was categorized into never smoking (smoked fewer than 100 cigarettes in lifetime) or smoking history, with never smoking considered a healthy level. Alcohol use was defined by daily consumption of alcohol, and a healthy level was defined as daily consumption of no more than one drink for women and no more than two drinks for men.^38^ Physical activity was assessed weekly by metabolic equivalent hours of leisure time (MET). MET was categorized into tertiles, and we considered participants in the top tertile to show evidence of adequate physical activity (healthy level).

Diet was characterized by the healthy eating index-2015 (HEI-2015).^39^ The HEI-2015 is a diet quality index to assess the adherence to 2015-2020 Dietary Guidelines for Americans.^39^ Intake of each HEI-2015 component was scored proportionately between the minimum and maximum standards. The details of HEI-2015 components and scoring standards are shown in **Table S1**. HEI-2015 scores range from 0 to 100. A healthy diet was defined as HEI-2015 scores in the top 40%, while those in the bottom 60% showed evidence of unhealthy eating.^40^

We constructed a composite healthy lifestyle score. For each lifestyle factor, we assigned 1 point for a healthy level and 0 point for an unhealthy level. The healthy lifestyle score was calculated by summing each individual lifestyle factor score, and possible scores ranged from 0 to 4. Higher healthy lifestyle scores indicated greater adherence to healthy behaviors. Healthy lifestyle scores were classified into approximate tertiles, comprising 0-1, 2, and 3-4 healthy behaviors.

### 4.4. SES assessment

The present study focused on individual-level SES, characterized by education, family income, occupation, health insurance, and food security.^41^ Education levels were categorized into less than high school, high school or equivalent, and college or above. Family income levels were defined as the ratios of family income to the poverty thresholds specific to the survey year. Family income levels were classified into low income (family income to poverty ratio ≤1), middle income (>1 and <4), and high income (≥4).^42^ Occupation was defined by each participant’s longest job, and categorized as blue collar (service, farming/forestry/fishing, precision production/craft/repair, and operator/fabricator/laborer), white collar (managerial/professional specialty, and technical/sales/administrative support), unemployed, or not in labor force (retired persons, homemakers, and students).^43^ Health insurance was classified into government insurance (Medicare, Medi-Gap, Medicaid, State Children’s Health Insurance Program, military health plan, Indian Health Service, state-sponsored health plan, or other government insurance), private insurance, both government and private insurance, or uninsured. Food insecurity was measured using 18-item Food Security Survey Module, and dichotomized into full food security and food insecurity.^44^

For composite SES, latent class analysis (LCA) was used to identify the presence of underlying constructs (or classes) in the observed SES components (education, occupation, income-to-poverty ratio, health insurance, and food security).^45^ LCA estimates conditional class membership probability and classifies individuals who are homogeneous in terms of particular criteria. We compared the performance of LCA with two, three, and four classes, and the optimal number of latent classes was determined based on the Akaike Information Criterion (AIC) and the Bayesian Information Criterion (BIC).^45^ We identified three latent classes - high, medium, and low SES groups.

### 4.5. Cognitive function assessment

Comprehensive cognitive testing was completed by participants aged ≥60 years during NHANES 2011-2012 and 2013-2014 cycles. Four tests were administered, including the Consortium to Establish a Registry for Alzheimer’s Disease (CERAD) Word Learning subtests to evaluate immediate and delayed learning ability,^46^ the Animal Fluency test to assess categorical verbal fluency (component of executive function),^47^ and the Digit Symbol Substitution Test (DSST) from the Wechsler Adult Intelligence Scale to assess processing speed, sustained attention, and working memory.^48^ Individual results of the four tests (CERAD immediate and delayed learning, Animal Fluency, DSST) converted to z-scores using age appropriate normal means. Individual test z scores are averaged to form the cognitive composite z-score, similar to previous studies.^49^

### 4.6. Covariates

Covariates were selected based on previous research and included age, sex, race/ethnicity, marital status, birthplace, and body mass index (BMI).^50^ Sex included male and female. Participants were assigned to one of these categories by NHANES based on their questionnaire responses. Race/ethnicity categories provided by NHANES included non-Hispanic White, non-Hispanic Black, non-Hispanic Asian, Hispanic, or other races including multiracial. Marital status included never married, married, or not married (including widowed, divorced, separated, or living with partner). Birthplace was categorized as born in or outside of the United States. BMI (kg/ m^2^) was computed as the ratio of weight (kg) and height squared (m^2^).

### 4.7. Statistical analyses

Mean and standard error (SE) were computed for continuous variables, and percentage (%) was calculated for categorical variables. Participant characteristics, composite cognition z-scores, and scores of the four individual cognitive tests were compared by composite healthy lifestyle and composite SES groups, using Chi-square or Fisher’s exact tests for categorical variables, and ANOVA or Kruskal-Wallis tests for continuous variables. The sampling weights and design variables were used for all analyses.

In the primary analysis, we utilized survey-weighted linear regression models to examine the associations of healthy lifestyle score and composite SES derived from LCA with composite cognition z-score, after adjusting for age, sex, race/ethnicity, marital status, and birthplace. To assess the joint associations of lifestyles and SES, we classified participants into nine groups based on three SES classes (low, medium, high) and three healthy lifestyle groups (0-1, 2, 3-4). Differences in cognition z-score and 95% confidence intervals were calculated with participants with low SES and 0-1 health lifestyle as the reference group. Likelihood-ratio test comparing models with and without the nine groups was used to test the interaction between healthy lifestyle score and composite SES.

In the secondary analysis of individual SES and lifestyle factors, we performed linear regression models to estimate differences in composite cognitive z-score and 95% confidence intervals. Model 1 included age, sex, race/ethnicity, marital status, birthplace, and individual lifestyle factors (smoking status, alcohol consumption, physical activity, and HEI-2015). Model 2 included age, sex, race/ethnicity, marital status, birthplace, and individual SES factors (education, family income, occupation, health insurance, and food security). Model 3 included age, sex, race/ethnicity, marital status, birthplace, as well as both individual lifestyle and SES components.

To test the robustness of our findings, we conducted several sensitivity analyses. First, to assess the different cognitive domains, we determined the relationships between lifestyle and SES factors with z-scores of each of the four cognitive test scores. Second, to assess potential differential influences between healthy lifestyle factors on cognition, we computed a weighted healthy lifestyle score. The weighted healthy lifestyle score was constructed as the sum of lifestyle factor scores where weights are beta coefficients of each individual lifestyle factor derived from Model 1 in the analysis of individual lifestyle factors. We then used this weighted healthy lifestyle score in a multivariable regression analysis. Furthermore, BMI could be an intermediate factor linking exposures and cognition, so we did not adjust for BMI in our primary analyses. However, in sensitivity analyses, we additionally controlled for BMI. Moreover, we repeated our analyses in a subpopulation excluding participants with stroke or cardiovascular disease, which might impair cognitive functions.^51^ Finally, to explore the impact of missing values on the observed results, we conducted multiple imputations with chain equation to impute missing values.^52^ This procedure used PROC MI in SAS to create 20 datasets for missing values and computed pooled effect estimates using PROC MIANALYZE. We repeated Models 1-3 on each of the imputed datasets and compared the effect estimates to the primary findings using measured data. All statistical analyses were performed using SAS (version 9.4, SAS Institute Inc.). Statistical significance was set at a two-sided *P*<0.05.

## Data Availability

All data and materials have been made publicly available at the National Center for Health Statistics website (https://www.cdc.gov/nchs/nhanes/index.htm).

## Abbreviations

AIC: Akaike Information Criterion
BIC: Bayesian Information Criterion
BMI: body mass index
CERAD: Consortium to Establish a Registry for Alzheimer’s Disease
DSST: Digit Symbol Substitution Test
FINGER: Finnish Geriatric Intervention Study to Prevent Cognitive Impairment and Disability
HEI: healthy eating index
LCA: latent class analysis
MAPT: French Multidomain Alzheimer Preventive Trial
MET: metabolic equivalent hours of leisure time
NHANES: National Health and Nutrition Examination Survey
PreDIVA: Dutch Prevention of Dementia by Intensive Vascular Care
SES: socioeconomic status.

## Acknowledgement

The authors would also like to thank the NHANES participants and the staff members for their contribution to data collection and for making the data publicly available.

## Author contributions

X.W. is the guarantor of this work and had full access to all the data in the study and takes responsibility for the contents of the manuscript. X.W. designed the study, conducted data analysis, and wrote the manuscript. K.M.B., H.L.P., R.L.A., and S.K.P. were involved in the design of the analysis plan, contributed to interpretation of the data, and critically revised the manuscript. All authors read and approved the final version of the paper.

