## Supplemental Material for "Associations of Healthy Lifestyle and Socioeconomic Status with Cognitive Function in U.S. Older Adults"

**Table S1** The healthy eating index-2015 components and scoring standards.*

| Component | Max points | Standard for maximum score | Standard for minimum score |
|---|---|---|---|
| Adequacy | | | |
| Total fruits (including fruit juice) | 5 | ≥0.8 cup equiv. per 1,000 kcal | No Fruits |
| Whole fruits (except fruit juice) | 5 | ≥0.4 cup equiv. per 1,000 kcal | No Whole Fruits |
| Total vegetables | 5 | ≥1.1 cup equiv. per 1,000 kcal | No Vegetables |
| Greens and beans | 5 | ≥0.2 cup equiv. per 1,000 kcal | No Greens and Beans |
| Whole grains | 10 | ≥1.5 oz equiv. per 1,000 kcal | No Whole Grains |
| Dairy | 10 | ≥1.3 cup equiv. per 1,000 kcal | No Dairy |
| Total protein foods | 5 | ≥2.5 oz equiv. per 1,000 kcal | No Protein Foods |
| Seafood and plant proteins | 5 | ≥0.8 oz equiv. per 1,000 kcal | No Seafood or Plant Proteins |
| Fatty acids† | 10 | (PUFAs + MUFAs)/SFAs ≥2.5 | (PUFAs + MUFAs)/SFAs ≤1.2 |
| Moderation | | | |
| Refined grains | 10 | ≤1.8 oz equiv. per 1,000 kcal | ≥4.3 oz equiv. per 1,000 kcal |
| Sodium | 10 | ≤1.1 gram per 1,000 kcal | ≥2.0 grams per 1,000 kcal |
| Added sugars | 10 | ≤6.5% of energy | ≥26% of energy |
| Saturated fats | 10 | ≤8% of energy | ≥16% of energy |

* The table was derived from Krebs-Smith SM, Pannucci TRE, Subar AF, et al. Update of the Healthy Eating Index: HEI-2015. J Acad Nutr Diet. Elsevier; 2018;118:1591–1602.

† Fatty acids include polyunsaturated fatty acids (PUFAs), monounsaturated fatty acids (MUFAs), and saturated fatty acids (SFAs).

**Table S2** The results from the latent class analysis for composite socioeconomic status (SES).

| | High SES | Medium SES | Low SES |
|---|---|---|---|
| | Probability | Probability | Probability |
| Income-to-poverty ratio | | | |
| Low (≤1) | 0.03 | 0.01 | 0.46 |
| Medium (>1 and <4) | 0.47 | 0.81 | 0.54 |
| High (≥4) | 0.5 | 0.18 | 0 |
| Education | | | |
| <high school | 0 | 0.29 | 0.46 |
| High school | 0.11 | 0.38 | 0.24 |
| College or higher | 0.89 | 0.33 | 0.3 |
| Occupation[*] | | | |
| Blue collar | 0.06 | 0.66 | 0.65 |
| White collar | 0.93 | 0.31 | 0.29 |
| Not in labor | 0.01 | 0.03 | 0.05 |
| Unemployed | 0 | 0 | 0.01 |
| Health insurance | | | |
| Uninsured | 0.03 | 0.03 | 0.18 |
| Government insurance | 0.24 | 0.35 | 0.65 |
| Private insurance | 0.29 | 0.21 | 0.06 |
| Both government and private | 0.44 | 0.41 | 0.11 |
| Food security | | | |
| Having food insecurity | 0.03 | 0.08 | 0.54 |
| Full food security | 0.97 | 0.92 | 0.46 |

[*] Blue collar occupations included service, farming/forestry/fishing, precision production/craft/repair, and operator/fabricator/laborer occupations. White collar occupations included managerial/professional specialty, and technical/sales/administrative support occupations. Not in labor force included retired persons, homemakers, and students.

**Table S3** Survey-weighted differences and 95% confidence intervals (95% CIs)* in z-scores of four cognitive tests with socioeconomic status and healthy lifestyle from the U.S. National Health and Nutrition Examination Survey 2011-2014.

| | CERAD† Word List learning | CERAD delayed recall | Animal Fluency | Digit Symbol Substitution Test |
|---|---|---|---|---|
| | Change (95% CI) | Change (95% CI) | Change (95% CI) | Change (95% CI) |
| Healthy lifestyle score | | | | |
| 0 or 1 healthy behavior | REF | REF | REF | REF |
| 2 healthy behaviors | 0.06 (-0.13, 0.25) | -0.03 (-0.16, 0.11) | 0.02 (-0.12, 0.15) | 0.07 (-0.08, 0.22) |
| 3 or 4 healthy behaviors | 0.12 (-0.05, 0.29) | 0.09 (-0.05, 0.23) | -0.03 (-0.18, 0.12) | 0.12 (-0.02, 0.25) |
| Socioeconomic status | | | | |
| Low SES | REF | REF | REF | REF |
| Medium SES | 0.07 (-0.11, 0.25) | -0.007 (-0.16, 0.10) | 0.13 (-0.07, 0.33) | 0.34 (0.16, 0.51)** |
| High SES | 0.45 (0.28, 0.62)*** | -0.03 (-0.16, 0.10) | 0.36 (0.20, 0.53)*** | 0.72 (0.55, 0.89)*** |

* Model was adjusted for age, sex, race/ethnicity, marital status, and birthplace.

† Consortium to Establish a Registry for Alzheimer's Disease (CERAD).

*P<0.05, **P<0.01, ***P<0.0001

**Table S4** Survey-weighted differences and 95% confidence intervals (95% CIs)* in cognitive composite z-score with socioeconomic status and weighted healthy lifestyle from the U.S. National Health and Nutrition Examination Survey 2011-2014.

| | Change (95% CI) |
|---|---|
| Weighted healthy lifestyle score | |
| Tertile 1 | REF |
| Tertile 2 | 0.01 (-0.06, 0.09) |
| Tertile 3 | 0.08 (0.03, 0.12)** |
| Socioeconomic status | |
| Low SES | REF |
| Medium SES | 0.13 (0.03, 0.23)* |
| High SES | 0.37 (0.28, 0.46)*** |

* Model was adjusted for age, sex, race/ethnicity, marital status, and birthplace.
*P<0.05, **P<0.01, ***P<0.0001

**Table S5** Survey-weighted differences and 95% confidence intervals (95% CIs)* in cognitive composite z-score with socioeconomic status and healthy lifestyle with an additional adjustment for body mass index from the U.S. National Health and Nutrition Examination Survey 2011-2014.

| | Change (95% CI) |
|---|---|
| Healthy lifestyle score | |
| Tertile 1 | REF |
| Tertile 2 | 0.03 (-0.05, 0.11) |
| Tertile 3 | 0.07 (0.005, 0.14)* |
| Socioeconomic status | |
| Low SES | REF |
| Medium SES | 0.13 (0.03, 0.24)* |
| High SES | 0.37 (0.29, 0.46)*** |

* Model was adjusted for age, sex, race/ethnicity, marital status, and birthplace.
*P<0.05, **P<0.01, ***P<0.0001

**Table S6** Survey-weighted differences and 95% confidence intervals (95% CIs)* in cognitive composite z-score with socioeconomic status and healthy lifestyle from the U.S. National Health and Nutrition Examination Survey 2011-2014 after excluding 283 participants with cardiovascular disease or stroke.

| | Change (95% CI) |
|---|---|
| Healthy lifestyle score | |
| 0 or 1 healthy behavior | REF |
| 2 healthy behaviors | 0.02 (-0.06, 0.10) |
| 3 or 4 healthy behaviors | 0.09 (0.01, 0.17)* |
| Socioeconomic status | |
| Low SES | REF |
| Medium SES | 0.13 (0.04, 0.22)** |
| High SES | 0.41 (0.32, 0.50)*** |

* Model was adjusted for age, sex, race/ethnicity, marital status, and birthplace.
*P<0.05, **P<0.01, ***P<0.0001

**Table S7** Survey-weighted differences and 95% confidence intervals (95% CIs)* in cognitive composite z-score with an additional interaction term† between socioeconomic status (SES) and healthy lifestyle from the U.S. National Health and Nutrition Examination Survey after excluding 283 participants with cardiovascular disease or stroke.

| | Socioeconomic status | | |
|---|---|---|---|
| Healthy lifestyle score | Low SES | Medium SES | High SES |
| 0 or 1 healthy behavior | REF | 0.22 (0.06, 0.38)** | 0.32 (0.16, 0.48)** |
| 2 healthy behaviors | -0.01 (-0.17, 0.16) | 0.04 (-0.11, 0.19) | 0.45 (0.32, 0.57)*** |
| 3 or 4 healthy behaviors | 0.05 (-0.08, 0.19) | 0.19 (0.04, 0.35)* | 0.49 (0.36, 0.60)*** |

* Model was adjusted for age, sex, race/ethnicity, marital status, and birthplace.

† P for interaction between SES and healthy lifestyle was 0.09.

*P<0.05, **P<0.01, ***P<0.0001

**Table S8** Survey-weighted differences and 95% confidence intervals (95% CIs) in cognitive composite z-score from linear regression models with 20 multiple imputations from the U.S. National Health and Nutrition Examination Survey 2011-2014.

| | Model 1 | Model 2 | Model 3 |
|---|---|---|---|
| | Change (95% CI) | Change (95% CI) | Change (95% CI) |
| Lifestyle factors | | | |
| Smoking status | | | |
| Former or current smoker | REF | – | REF |
| Never smoker | 0.08 (0.01, 0.14)* | – | 0.02 (-0.03, 0.08) |
| Alcohol consumption[*] | | | |
| High | REF | – | REF |
| Low or moderate | -0.14 (-0.16, -0.12)** | – | -0.07 (-0.16, 0.007) |
| Physical activity | | | |
| Not healthy | REF | – | REF |
| Healthy | 0.05 (-0.01, 0.11) | – | 0.05 (0, 0.10)* |
| Healthy eating index | | | |
| Not healthy | REF | – | REF |
| Healthy | 0.12 (0.11, 0.13)*** | – | 0.06 (0.02, 0.10)** |
| Socioeconomic status | | | |
| Poverty-to-income ratio | | | |
| ≤1 | – | REF | REF |
| >1 and <4 | – | 0.08 (0.005, 0.15)* | 0.07 (0.0007, 0.15)* |
| ≥4 | – | 0.17 (0.10, 0.25)*** | 0.16 (0.08, 0.24)*** |
| Education | | | |
| <High school | – | REF | REF |
| High school or equivalent | – | 0.12 (0.07, 0.17)*** | 0.11 (0.06, 0.17)*** |
| >High school | – | 0.32 (0.25, 0.38)*** | 0.30 (0.23, 0.37)*** |
| Insurance | | | |
| Uninsured | – | REF | REF |
| Covered by government | – | 0.06 (-0.03, 0.14) | 0.06 (-0.03, 0.15) |
| Covered by private | – | 0.07 (-0.02, 0.17) | 0.06 (-0.03, 0.16) |
| Covered by both | – | 0.06 (-0.02, 0.14) | 0.06 (-0.03, 0.14) |
| Food security | | | |

| | | | |
|---|---|---|---|
| Having food insecurity | – | REF | REF |
| Full food security | – | 0.09 (0.02, 0.17)* | 0.09 (0.02, 0.16)* |
| Occupation | | | |
| Blue collar† | – | REF | REF |
| White collar‡ | – | 0.18 (0.12, 0.24)*** | 0.17 (0.11, 0.23)*** |
| Not in labor force§ | – | 0.06 (-0.05, 0.16) | 0.05 (-0.06, 0.16) |
| Unemployed | – | 0.27 (0.04, 0.49)* | 0.28 (0.05, 0.51)* |

* Low alcohol consumption was defined as daily consumption of one drink or fewer for women and two drinkers or fewer for men, and otherwise coded as moderate or high consumption.

† Blue collar occupations included service, farming/forestry/fishing, precision production/craft/repair, and operator/fabricator/laborer occupations.

‡White collar occupations included managerial/professional specialty, and technical/sales/administrative support occupations.

§ Not in labor force included retired persons, homemakers, and students.

*P<0.05, **P<0.01, ***P<0.0001

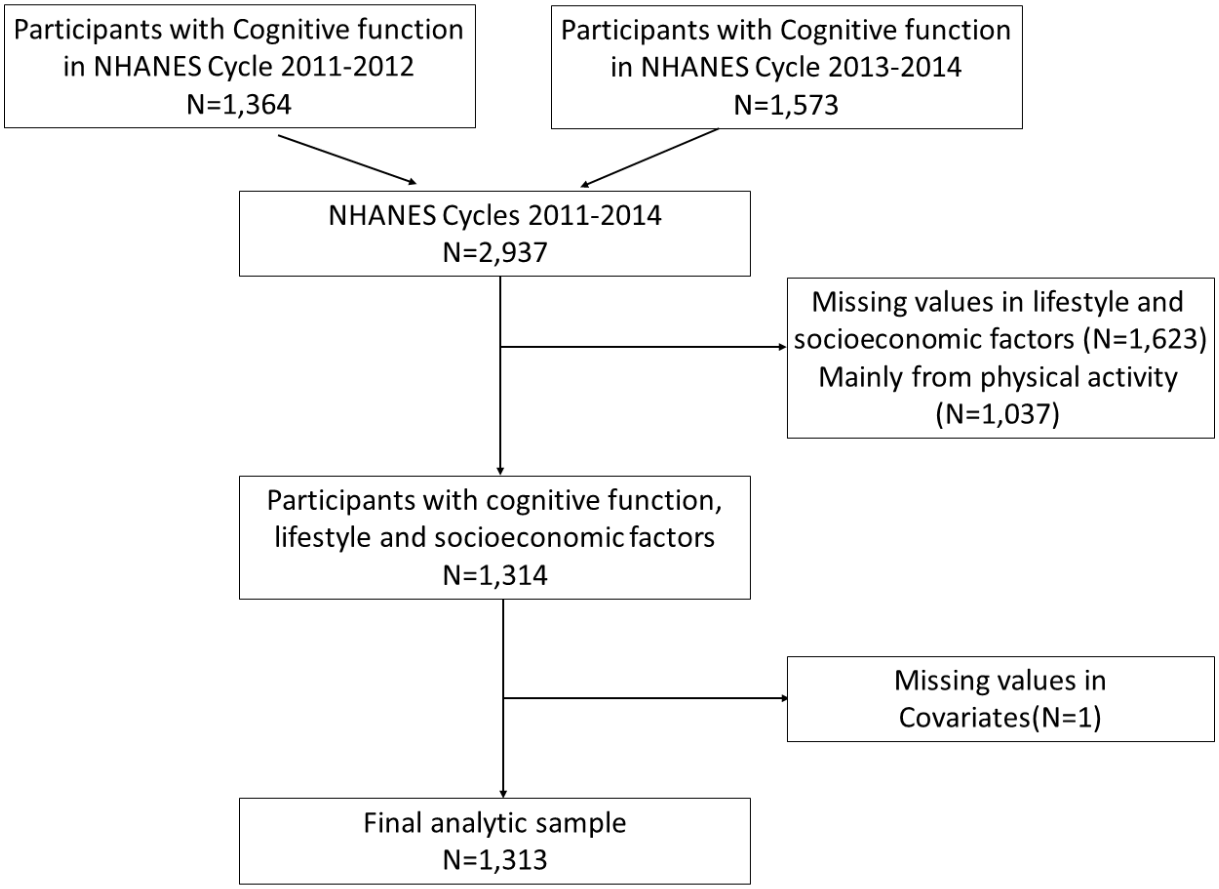


**Figure S1** The flowchart of participant selection in the present study.
